# Estimating controlled direct treatment effects on pain intensity using structural mean models: application to pain randomized controlled trials

**DOI:** 10.1101/2025.04.09.25325306

**Authors:** Rui Wang, Patrick J. Heagerty, Kwun Chuen Gary Chan, Pradeep Suri

**Affiliations:** Department of Biostatistics, University of Washington, Seattle, USA; Clinical Learning, Evidence, and Research (CLEAR) Center, University of Washington, Seattle, USA; Seattle Epidemiologic Research and Information Center, VA Puget Sound Health Care System, Seattle, USA; Division of Rehabilitation Care Services, VA Puget Sound Health Care System, USA; Department of Rehabilitation Medicine, University of Washington, Seattle, USA

## Abstract

Analytical methods to incorporate potential concurrent analgesic use into primary statistical summaries are underutilized in pain randomized controlled trials (RCTs). Without valid inclusion of analgesic use, the primary analyses of pain may produce diminished estimated treatment effects. We used contemporary causal inference methods that can account for concurrent treatment to reanalyze RCT data examining the effect of epidural steroid injection (ESI). Specifically, we define an “attributable to ESI estimand”, which is the controlled direct effect of ESI. We used a simple composite pain intensity outcome, the QPAC_1.5,_ and structural mean models (SMM) to estimate the target estimand. Compared to traditional methods such as strict intention to treat analysis (strict ITT), SMMs can account for analgesic use without assuming sequential ignorability. We estimated treatment effects of ESI on leg pain intensity (LPI) using the numeric rating scale (NRS) with strict ITT, 3 SMM estimating methods (estimating equations [EE], g-estimation, and generalized method of moments [GMM]), and the QPAC_1.5_. The treatment effect of ESI on LPI using strict ITT was -0.751 NRS points (95% confidence interval [CI]: [-1.287, -0.214]). Estimates for the attributable to ESI estimand were -0.864 (95% CI: [-3.207, 1.478]) for EE, -0.935 (95% CI: [-1.779, 0.090]) for g-estimation, -0.653 (95% CI: [-1.218, -0.089]]) for GMM, and -0.930 (95% CI: [-1.508, - 0.352]) for the QPAC1.5. We illustrate how contemporary causal inference methods and alternative estimands can be used to account for concurrent analgesic use in pain RCTs, in a manner that may result in larger treatment effects.

## 1. Introduction

Randomized controlled trials (RCT) are widely used in evaluating new pain treatments. Due to ethical considerations, as-needed pain medication use is typically allowed in pain RCTs [3; 5]. At the same time, the effect size of intercurrent analgesics may equal or exceed that of the randomized treatments being studied in pain randomized controlled trials.[22] These aspects taken together may contribute to the fact that most pain treatments have only small treatment effects as estimated using intention-to-treat analysis. Recent FDA guidance explicitly recognizes potential problems posed by intercurrent treatments such as analgesic use. [10] However, intercurrent analgesic use is often ignored in pragmatic RCTs or dealt with using analytic approaches (e.g. inclusion of analgesic use in a binary “responder outcome”) that can reduce power or impart bias [21], or using design features (e.g. strict study-mandated rescue medication use) that can encourage participant drop-out and lead to biased estimates of treatment effect [17]. Importantly, analgesic use may be a mediator between pain intensity at one time point and pain intensity at another time point, so that adjustment for analgesic use as a covariate is generally not appropriate. [21] Formal statistical methods are needed to guide the development of strategies that can account for intercurrent analgesic use.

Our team has proposed simple methods and novel measures to account for intercurrent analgesic use that might improve power and accuracy in pain RCTs. [21; 22] Separately, existing and well-established biostatistical methods from the causal mediation analysis literature can address the problem of whether and how to account for intercurrent analgesic use, [8; 11; 23] especially when there is unmeasured confounding between intercurrent analgesic use and pain intensity outcomes. However, such methods to our knowledge are not used in the vast majority of pain RCTs. [9] A recent consensus statement on pragmatic pain RCT methods states “Proper adjustment for such effects, when warranted, requires the use of sophisticated causal inference methods”([9], page 7), but provides no examples of or guidance on *which* causal inference methods to use, or how they may achieve (or fail to attain) the goal of accounting for intercurrent analgesic use.

Given that certain appropriate causal inference methods were developed decades ago and have been used with increasing frequency recently, we considered that the delay in their uptake in pain RCTs may be due to lack of familiarity with their goals and assumptions among pain researchers, and questions about how best to use such methods. Therefore, the current study sought to illustrate the use of such methods to address intercurrent analgesic use in a pain RCT, with an emphasis on one type of contemporary causal inference method, structural mean models (SMMs). The aim of this study was to illustrate the use of contemporary causal inference methods, including but not limited to structural mean models, to account for intercurrent analgesic in an RCT examining the treatment effect of epidural steroid injections (ESI) on pain intensity, among patients with lumbar spinal stenosis. [6]

## 2. Methods

This study analyzed data from the Lumbar Epidural Steroid Injections for Spinal Stenosis (LESS) trial, the largest double-blind randomized controlled trial of a percutaneous, non-surgical procedure for lumbar spinal disorders conducted to date. [6] To allow the work to be broadly accessible to readers, the text of this manuscript contains basic information about methodology but relegates most technical biostatistical details to the Supplemental Methods.

### 2.1 Conceptual overview

In a pain RCT, the effect on pain intensity of treatment versus control can be estimated using a standard difference in mean estimator or regression-based estimator due to the randomization of treatment assignment.[13] The effect of treatment assignment is also called the intention to treat (ITT) effect. However, the existence of intercurrent analgesic use may partially nullify the direct effect on pain intensity (Figure 1). Accordingly, the outcome of greater interest may be the controlled direct effect (CDE) of treatment assignment on pain intensity, which represents the average effect of treatment versus control if all participants in the study sample hadn’t taken intercurrent analgesics at the primary pain intensity endpoint. To formally identify the CDE, a sequential ignorability assumption is usually needed. [8] This assumption has two implications: first, the causal relationship between treatment assignment and outcome is unconfounded by unobservable baseline variables; second, the causal relationship between the mediator (intercurrent analgesic use) and outcome is unconfounded by unobservable baseline variables. While the former assumption holds by study design in an RCT, the second one may be hard to justify even in an RCT (since post-randomization intercurrent analgesic use is not randomized), as the causal relationship between mediator (intercurrent analgesic use) and outcome may be confounded by unobserved underlying pain intensity or other unmeasured factors.

**Figure 1.**
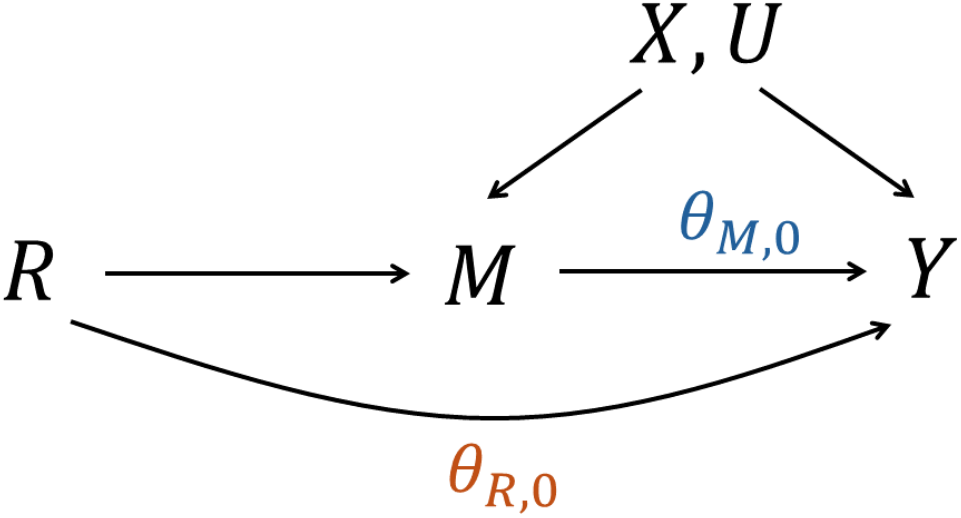
Causal diagram of LESS trial. Here R denotes the randomized treatment assignment, M denotes the analgesic use, X denoted the observed baseline confounders, U denotes the unmeasured confounder and Y denotes outcome. *θ*_*M*,0_ is the effect of analgesic use on outcome, *θ*_*R*,0_ is the direct effect of randomized treatment assignment on the outcome.

A rank preserving model (RPM) accompanied by g-estimation has been proposed to address the issue of unmeasured confounding by a mediator (intercurrent analgesic use) affecting the outcome variable (Table 1). [23] The RPM relaxes the sequential ignorability assumption while it imposes stronger assumptions on effect homogeneity. In this case, interactions between randomized treatment assignment and baseline covariates could serve as instrumental variables.[23] G-estimation can be used for estimation in such cases when instruments are constructed in such a way. [23] In our paper, we use SMMs instead of RPMs to parameterize the causal effects that we are interested in. SMMs are closely related to RPMs, however, the assumption involved in SMMs is weaker than those in RPMs. We recommend readers consult chapter 14.4 of [8] for a more detailed discussion on the differences between RPM and SMM. Roughly speaking, RPMs involve an additional untestable assumption that the error distribution is the same across different levels of treatment and analgesic usage. We also tried to leverage the fact that interactions between all baseline covariates and randomized treatment assignment could serve as instrumental variables. [18; 23] Using more instruments could potentially reduce the bias and improve the efficiency of estimation. [7] Therefore, we also use generalized method of moments (GMM), [7] which allow us to use multiple instruments to estimate the direct effect of randomized treatment assignment (Table 1). However, it should be noted that the GMM is not necessarily better than g-estimation when the sample size is not large and when some instruments are weak, meaning that their correlation with analgesic use is small.

**Table 1.** Comparison of different methods of analysis with detail on the goal of the approach through statement of the estimand, and details on the specific strategies.

| Methods | Descriptions | Estimands |
| --- | --- | --- |
| ITT | • Ignores intercurrent analgesic use.<br>• Directly regresses outcomes on treatment assignment, adjusting or not adjusting covariates. | *Strict ITT estimand*<br>(Total effect of |
|  |  | randomized treatment assignment) |
| Baron/Kenny | • Accounts for intercurrent analgesic use.<br>• Directly regresses outcome on analgesic use, treatment assignment and other covariates.<br>• The coefficient of analgesic use can be interpreted as the direct effect of treatment.<br>• Assumes no unmeasured confounding between analgesic use and outcomes. | The *attributable to ESI* *estimand* and the *effect of intercurrent analgesic use on outcomes* |
| SMM | • Accounts for intercurrent analgesic use.<br>• Doesn't assume no unmeasured confounding between analgesic use and outcomes.<br>• Assumes baseline covariates are not effect-modifiers.<br>• Multiple estimation methods can be applied, including estimating equations, g-estimation and generalized method of moments. All the estimating methods are based on using the interactions of baseline covariates and randomized treatment assignment to construct instrumental variables.<br>○ The EE method uses one baseline covariate and randomized treatment assignment to construct an instrumental variable.<br>○ The g-estimation uses one baseline covariate and randomized treatment assignment to construct an instrumental variable.<br>Furthermore, g-estimation involves fitting | The *attributable to ESI* *estimand* and the *effect of intercurrent analgesic use on outcomes* |
|  | another nuisance model for the outcome without analgesic use, which could improve estimation efficiency in practice.<br>○ GMM (Generalized method of moments) uses multiple baseline covariates and randomized treatment assignment to construct instrumental variables. Furthermore, GMM involves fitting two other nuisance models for the outcome and analgesic use, which could improve estimation efficiency in practice. |  |
| QPAC<sub>1.5</sub> | • Accounts for intercurrent analgesic use.<br>• Doesn't assume no unmeasured confounding between analgesic use and outcomes.<br>• Assumes the effect of intercurrent analgesic use is homogeneous and known. However, the value assumed for the effect intercurrent analgesic use can be varied depending on what is known regarding the target population. | The *attributable to ESI estimand* |

Abbreviations: ITT analysis = Intention to treat analysis, SMM = structural mean models, QPAC<sub>1.5</sub>= quantitative pain and analgesia composite outcome based on adding 1.5 points to pain intensity for those who were taking an analgesic, EE = estimating equation, ESI = epidural steroid injection.

We have previously reported that the treatment effect of epidural steroid injection (ESI) on leg pain intensity NRS at 3 weeks post-randomization using strict ITT was estimated as -0.6 NRS points (95% CI [-1.2, -0.1]) .[6] In this paper, we focused on leg pain intensity (LPI) as well as back pain intensity (BPI) to allow comparability with our prior work on intercurrent treatment, and because the primary target of analgesics is pain intensity, so that while effects of analgesics on other pain domains may exist, they are generally thought to occur secondary to changes in pain intensity. Prior to the start of analyses, we decided to focus on treatment effects at 3 weeks because (1) only the 3-week comparison in the LESS trial was truly randomized (as treatments were allowed to be repeated immediately after the 3-week assessment, depending on the participant’s choice and not in a randomized manner), and (2) the effect of corticosteroids on pain outcomes is generally thought to be short-lived (peaking in 1-3 weeks, and decreasing thereafter). The primary ITT analysis in the LESS study ignored analgesic use (as do most pragmatic pain RCTs). [6] This “original” analysis in LESS targeted an estimand that is the difference in mean LPI and BPI for all randomized subjects at 3-week follow-up, irrespective of whether the difference is attributable to ESI or whether it is potentially impacted by concurrent analgesic use, which we refer to henceforth as the ‘strict ITT estimand’. In the current study, we used SMM to analyze the treatment effect of ESI on 3-week LPI and BPI NRS outcomes, targeting an alternative estimand, which we refer to here as “the difference in mean leg pain intensity for all randomized subjects at 3-week follow-up that is attributable to ESI (the initially randomized treatment) and not affected by concurrent analgesic use at the time of the primary endpoint”. For simplicity we refer to this henceforth as “the attributable to ESI estimand”. The attributable to ESI estimand is a specific example of what we term the “attributable to randomized treatment estimand” in a pain RCT.

### 2.2 Study sample and assessments

The LESS trial was a double-blind, randomized, controlled trial, which involved 400 participants aged 50 years or older with moderate-to-severe leg pain recruited from 16 sites in United States between April 2011 and August 2013. Participants had symptoms of neurogenic claudication and lumbar central canal spinal stenosis on magnetic resonance imaging or computed tomography. Other inclusion criteria included an average pain rating of more than 4 and a score of 7 or higher on RMDQ. Demographic, clinical and disease-history information, and expectations of pain relief after receiving an injection were measured at baseline. Participants were randomized to receive ESI with glucocorticoids plus lidocaine (the “steroid”group) or an epidural injection with lidocaine alone. Outcomes were measured at 1, 2, 3 and 6 weeks after randomization through phone, in-person reviews or mailed questionnaires.

### 2.3 Baseline measures

Baseline characteristics including sociodemographic (age, sex, ethnicity, race, current employment status, education, marital status), body mass index, smoking status, baseline analgesic use, LPI and BPI were recorded.

### 2.4 Analgesic use

Concurrent analgesic use was measured every week by in-person reviews or mailed questionnaires. Participants were asked about their daily dose of opioid medication and non-opioid medication. In this paper, analgesic use is defined as a binary variable with a value of 1 if there was any analgesic use (including opioid medication and/or non-opioid medication) in week 3, and a value of 0 if no analgesic was reported.

### 2.5 Outcome measures

Outcome measures included LPI and BPI. LPI reflected average leg pain intensity on an 11-point numerical rating scale (NRS) in the past week. BPI reflected average back pain intensity on an 11-point NRS in the past week.

### 2.6 Statistical analysis

#### 2.6.1. Descriptive statistics

Summary statistics for baseline characteristics and primary outcomes were described by arms.

#### 2.6.2. Intention to treat (ITT) analysis

A “strict ITT” analysis was used to evaluate the effectiveness of treatment assignment. Two regression-based methods were used for estimating the ITT effect: (1) regressing outcome on treatment assignment, recruitment site and baseline values of both outcome measures (method 1) as was done in the original trial [6], and (2) regressing outcome on treatment assignment and all other baseline covariates (method 2) for potential improvement of efficiency. [4] Heteroskedasticity-robust standard error estimator was used for calculating standard errors [26] and constructing 95% confidence intervals.

#### 3.6.3. Traditional regression-based mediation analysis

Traditional regression-based mediation analysis was applied. [1] Let *R* be the baseline randomization (*R* = 1 refers to those who were randomized to receive glucocorticoids plus lidocaine, *R* = 0 refers to those who were randomized to receive lidocaine alone), *M* be the binary concurrent analgesic use, *Y* be the outcome measure (leg pain intensity or back pain intensity), *X* be covariates. The regression model is specified as

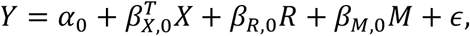

where the error term *ϵ* is assumed to be independent of *X,R* and *M*, which corresponds to the case where there is no unmeasured confounding between *M* and *Y*. Here *β*_*R*,0_ can be interpreted as the direct effect of randomization on leg pain intensity, *β*_*M*,0_ can be interpreted as the effect of analgesic use on leg pain intensity.

#### 2.6.4. QPAC_1.5_

We applied the QPAC_1.5_ method previously proposed in [21]. The QPAC_1.5_ is a simple method that combines information from concurrent analgesic use and pain intensity to create a composite outcome, resulting in a single value that reflects both pain intensity and concurrent analgesic use. It assumes a perceived beneficial effect on pain intensity of 1.5 NRS points among all analgesic users who have self-selected to use analgesics, based on an earlier study of people with low back pain. [22] As used in the current study, the QPAC_1.5_ targets the attributable to ESI estimand. A weakness of the QPAC_1.5_ is that, unlike more principled approaches such as SMMs, it assumes the same (beneficial) value for the effect of intercurrent analgesic use on pain intensity, without variability. Strengths of the approach are that it can be easily applied by non-biostatisticians without the more complex biostatistical analyses required by traditional regression-based mediation analysis and SMMs, and it is intuitive and easily understood. Additionally, while the QPAC_1.5_ assumes a beneficial 1.5-NRS-point among analgesic users, this value can be altered depending on what is known regarding the average perceived beneficial effect of analgesics on pain intensity among analgesic users in a given target population or study sample. [21]

We used two steps to apply the QPAC_1.5_ in this analysis. The first step was calculating the pain intensity-concurrent analgesic use composite outcome 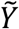 by 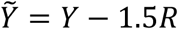. The second step was regressing 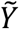 on randomized treatment assignment *R* and other covariates.

#### 2.6.5. Structural mean models

We adopt the potential outcome framework [16; 19] and use *Y*(*r,m)* to denote the potential outcome we would have observed if *R* were set to be *r* and *M* were set to be *m*. The SMM is specified as

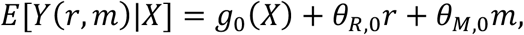

where *g*_0_(*X)* is an unspecified function of covariates. *θ*_*R*,0_ is the attributable to ESI estimand, which is also called controlled direct effect in causal inference literature [15]. *θ*_*M*,0_ is the effect of analgesic use on the outcome.

Additional assumptions are needed for identifying *θ*_*R*,0_ and *θ*_*M*,0_. First, the stable unit treatment value assumption (SUTVA) needs to be made, which means the potential outcomes for any participant do not vary with the treatments and analgesic use assigned to other participants, furthermore, there are no different forms or versions of each treatment and analgesic use level. [13] Second, we assume the randomization is independent of potential outcomes and baseline covariates; this assumption holds automatically by study design. In contrast to traditional Baron-Kenny-type methods, unmeasured confounding between analgesic use and the outcome is allowed since the actual analgesic use M does not appear as an independent variable in SMM.

Following Ten Have et al., [23] the moment conditions in SMMs can be constructed by treating interactions between randomization and baseline covariates as instrumental variables. In our paper, three different estimation methods were applied for estimating *θ*_*R*,0_ and *θ*_*M*,0_: estimating equations or EE, [24] g-estimation, [14; 25] and GMM[7]. Technical details can be found in the supplementary materials.

Different construction of instrumental variables could lead to different results. We centered and scaled the covariates when we constructed the instruments. For estimating equation and g-estimation approaches, we used the interaction between age and randomization as an instrument since both estimating equation and g-estimation approaches only allow for one instrument, and age had a relatively large correlation with analgesic use in the LESS data. For GMM, we used <u>all</u> the baseline covariates to construct instrumental variables and used an efficient weighting matrix. [7] The use of additional instruments and the efficient weighting matrix in GMM aims to improve estimation efficiency compared to the estimating equation and g-estimation approaches that use a single instrument. We also used the J-test to test if the moment conditions implied by the SMM were corrected specified.

## 3. Results

### 3.1 Baseline characteristics of the study participants

The LESS study included 400 participants, where 200 participants were assigned to the lidocaine alone group and glucocorticoids plus lidocaine were assigned to the glucocorticoids plus lidocaine group. There is missingness in those variables. After excluding samples with missing covariates, mediator or outcomes, the total number of samples we included for analysis is 358. As the proportion of missing data was low (∼10%), we performed complete-case analysis.

### 3.2 Results using intention to treat analysis

Table 3 shows the results for the “strict ITT” analysis. The estimated strict ITT treatment effect on LPI was -0.751(95% CI: [-1.287, -0.214]) in model 1 (adjusting for site and baseline covariates) and -0.782 (95% CI: [-1.336, -0.228]) in model 2 (adjusting for all baseline covariates). The estimated strict ITT treatment effect on BPI was -0.833 (95% CI: [-1.350, -0.316]) in model 1 and -0.932 (95% CI: [-1.465, -0.399]) in model 2. Compared to participants who were assigned to lidocaine alone, participants who were assigned to glucocorticoids plus lidocaine had lower LPI and BPI at week 3.

**Table 2.** Descriptive statistics. Continuous variables were presented as means (standard deviations) and categorical variables were presented as counts (proportions).

|  | Lidocaine alone | glucocorticoids plus lidocaine |
| --- | --- | --- |
| n | 200 | 200 |
| Age (mean (SD)) | 68.03 (10.16) | 67.97 (9.81) |
| BMI (mean (SD)) | 29.67 (6.01) | 31.09 (6.48) |
| Sex (%) |  |  |
| Female | 104 (52.0) | 117 (58.5) |
| Male | 96 (48.0) | 83 (41.5) |
| Race(%) |  |  |
| White | 139 (69.5) | 137 (69.2) |
| Black | 52 (26.0) | 53 (26.8) |
| Others | 9 ( 4.5) | 8 ( 4.0) |
| Education(%) |  |  |
| Less than college | 74 (37.0) | 65 (32.7) |
| College | 81 (40.5) | 99 (49.7) |
| Graduate School | 45 (22.5) | 35 (17.6) |
| Employment (%) |  |  |
| Currently no work | 126 (63.0) | 141 (70.5) |
| Full time or part time | 74 (37.0) | 59 (29.5) |
| Marital Status (%) |  |  |
| Married or living with |  |  |
| partner | 111 (55.8) | 126 (63.0) |
| Others | 88 (44.2) | 74 (37.0) |
| Smoking status(%) |  |  |
| Not a current smoker | 168 (84.0) | 174 (87.4) |
| Current smoker | 32 (16.0) | 25 (12.6) |
| BPI at baseline (mean (SD)) | 6.62 (2.61) | 6.70 (2.33) |
| BPI at week 3 (mean (SD)) | 4.60 (2.65) | 3.98 (2.66) |
| LPI at baseline (mean (SD)) | 6.62 (2.61) | 6.70 (2.33) |
| LPI at week 3 (mean (SD)) | 5.01 (2.81) | 4.38 (2.65) |
| Analgesic use* (w3, all) (%) |  |  |
| No | 92 (49.5) | 113 (59.8) |
| Yes | 94 (50.5) | 76 (40.2) |

Abbreviations: BMI=Body Mass Index, SD = standard deviation, w3 = week 3.
\*Either self-reported opioid or non-opioid medication

**Table 3.** Intention to treat analysis*.

|  | LPI |  |  | BPI |  |  |
| --- | --- | --- | --- | --- | --- | --- |
|  | Estimate | SE | 95% CI | Estimate | SE | 95% CI |
| Model 1 | -0.751 | 0.273 | [-1.287, -0.214] | -0.833 | 0.263 | [-1.350, -0.316] |
| Model 2 | -0.782 | 0.281 | [-1.336, -0.228] | -0.932 | 0.271 | [-1.465, -0.399] |

Abbreviations: LPI=leg pain intensity, BPI = back pain intensity, SE = standard error, CI = confidence interval.
\*Model 1 adjusts for site and baseline pain intensity. Model 2 adjusts for all baseline covariates.

### 3.3 Results using traditional regression-based mediation analysis

Table 4 shows the results for traditional regression-based mediation analysis. For LPI, the treatment effect estimate for the attributable to ESI estimand was -0.756 (95%: [-1.321, - 0.191]), which means that comparing the lidocaine alone group, LPI for participants who were assigned to glucocorticoids plus lidocaine was 0.756 lower if all participants hadn’t taken analgesics. This value for the treatment effect on LPI is generally similar in magnitude to that obtained for strict ITT using method 1 (−0.751) and method 2 (−0.782) from Table 2. Similarly, for BPI, the estimate is -0.875 (95%: [-1.415, -0.335]), which means that, compared to the lidocaine-alone group, BPI for participants who were assigned to glucocorticoids plus lidocaine was 0.875 lower, if all participants hadn’t taken analgesics. This value for the treatment effect on BPI is generally similar in magnitude to that obtained for strict ITT using method 1 (−0.833) and method 2 (−0.932) from Table 2. The direct effects of analgesic use were estimated as 0.262 (95% CI: [-0.374, 0.897]) for LPI and 0.579 (95% CI: [-0.036, 1.194]) for BPI. The point estimates are positive with 95% CIs including 0, suggesting a non-significant yet detrimental effect of analgesic use (i.e., that analgesics worsen pain intensity).

**Table 4.** Traditional regression-based mediation analysis*.

|  | LPI |  |  | BPI |  |  |
| --- | --- | --- | --- | --- | --- | --- |
|  | Estimate | SE | 95% CI | Estimate | SE | 95% CI |
| Direct effect of the<br>randomized treatment<br>(R) on the pain<br>intensity outcome (Y) | -0.756 | 0.287 | [-1.321, -0.191] | -0.875 | 0.274 | [-1.415, -0.335] |
| Direct effect of the<br>analgesic use mediator<br>(M) on the pain<br>intensity outcome (Y) | 0.262 | 0.323 | [-0.374, 0.897] | 0.579 | 0.313 | [-0.036, 1.194] |

Abbreviations: LPI=leg pain intensity, BPI = back pain intensity, SE = standard error, CI = confidence interval.
\*All baseline variables are adjusted for.

### 3.5 Results using the QPAC_1.5_

Table 5 shows the results using the QPAC_1.5_. The treatment effect estimates on pain intensity for the attributable to ESI estimand were -0.930 (95% CI: [-1.508, -0.352]) for LPI and -1.080 (95% CI: [-1.644, -0.516]) for BPI. These estimates of the attributable to ESI estimand using the QPAC_1.5_ are slightly bigger than the ITT estimates and from traditional regression-based mediation analysis estimates.

**Table 5.**
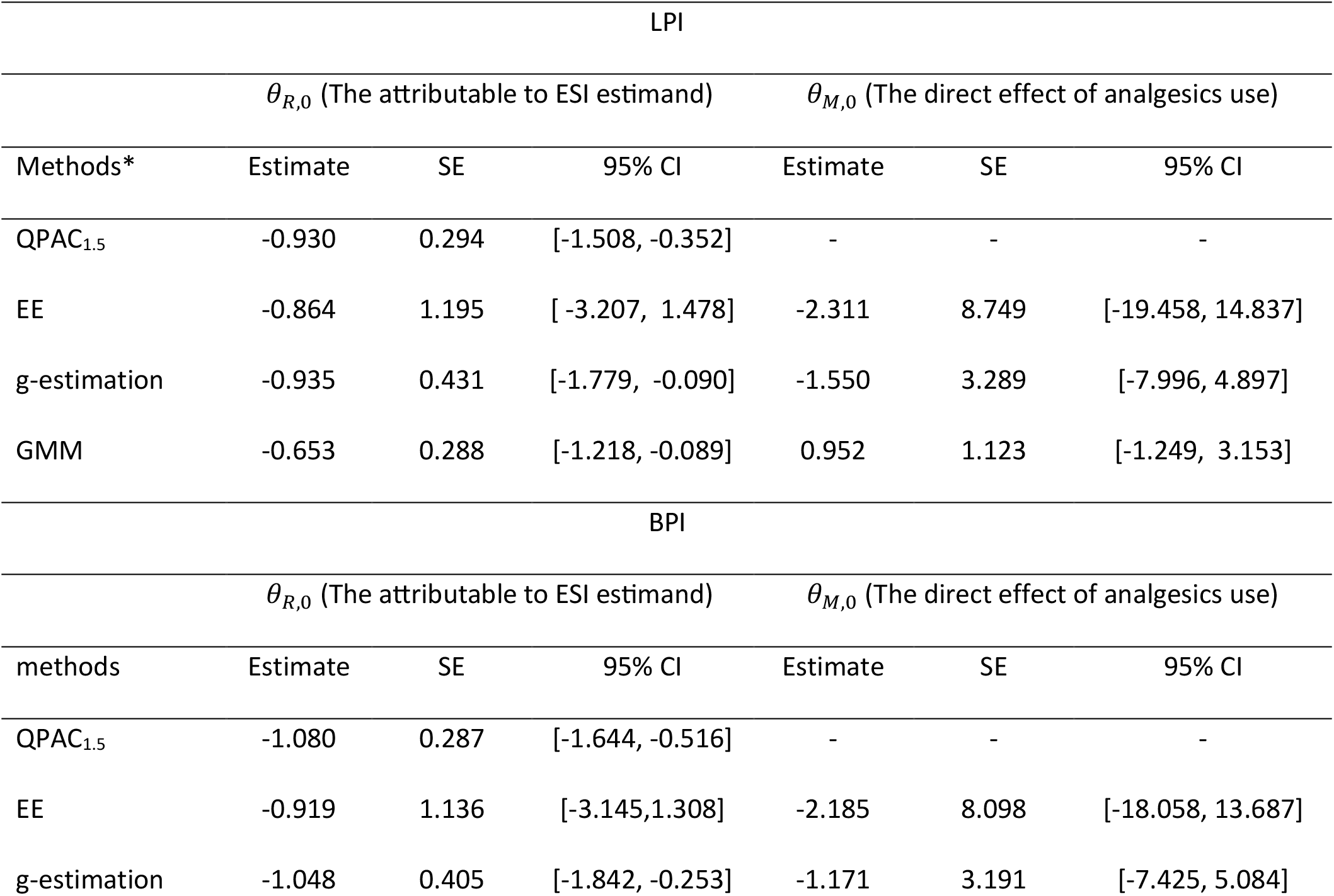
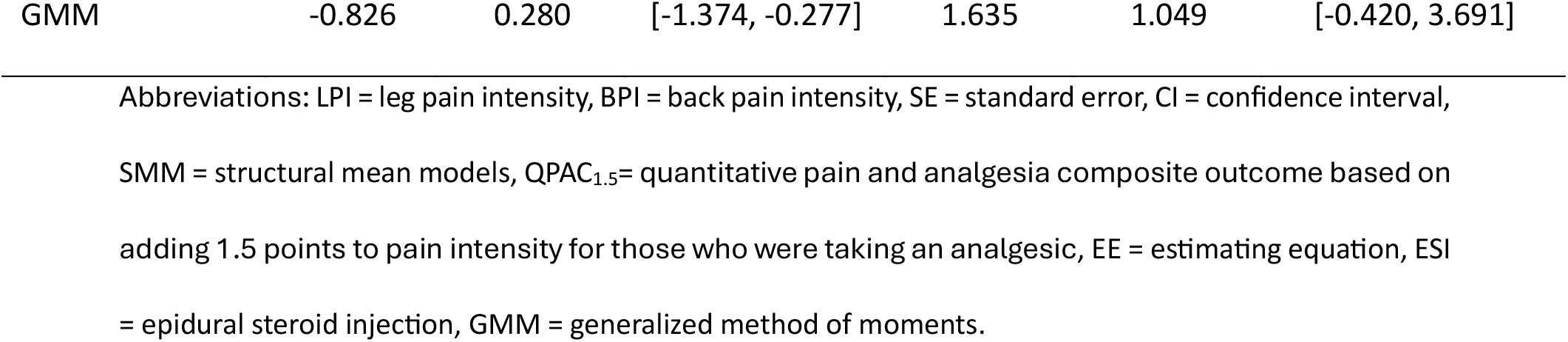
Analyses using the QPAC_1.5_ and SMMs.

### 3.4 Results using SMMs

Table 5 shows the results using SMM. For LPI, the treatment effect estimates for the attributable to ESI estimand were -0.864 (95% CI: [-3.207, 1.478]) for EE, -0.935 (95% CI: [-1.779, -0.090]) for g-estimation, and -0.653 (95% CI: [-1.218, -0.089]) for GMM. These treatment effect estimates with EE and g-estimation are slightly bigger than the ITT estimates and estimates from traditional regression-based mediation analysis. Interestingly, the estimate of the attributable to ESI estimand from g-estimation is very similar to the corresponding results of QPAC_1.5_ and the estimate of *θ*_*M*,0_ (−1.55) is very similar to the effect of intercurrent analgesic use assumed by the QPAC_1.5_ (−1.5 NRS points). The estimate of the attributable to ESI estimand from GMM is slightly smaller than ITT estimates and estimates from traditional regression-based mediation analysis. The estimate of the direct effect of analgesics from GMM is positive, with the 95% CI including zero. Compared to the estimating equation approach, g-estimation greatly shrank the standard error estimate (from 1.195 to 0.431), and GMM further shrank the standard error estimate to 0.288. Estimates for *θ*_*M*,0_ showed similar patterns, although the 95% CIs for *θ*_*M*,0_ were too wide to be informative. The p-value for the J-test was 0.443, which did not suggest evidence of model misspecification.

For BPI, the estimates for the attributable to ESI estimands are -0.919 (95% CI: [-3.145, 1.308]) for EE, -1.048 (95% CI: [-1.842, -0.253]) for g-estimation, and -0.826 (95% CI: [-1.374, -0.277]) for GMM. The overall patterns of the estimates are very similar to the LPI case: estimates of the attributable to ESI estimand from EE and g-estimation are slightly bigger than the ITT estimates and estimates from traditional regression-based mediation analysis. The estimate of the attributable to ESI estimand from g-estimation is very similar to the corresponding results using the QPAC_1.5_. The estimate of the attributable to ESI estimand from GMM is similar or slightly smaller than ITT estimates and estimates from traditional regression-based mediation analysis. The patterns for standard errors are also similar to those seen with the LPI estimates: g-estimation greatly shrinks the standard error estimate comparing to EE, and GMM further shrinks the standard error estimate comparing to g-estimation. The p-value for the J-test was 0.322, against any evidence of model misspecification.

## 4. Discussion

In this study, we applied contemporary causal inference statistical methodology to analyze pain RCT data while considering intercurrent analgesic use. We compared different contemporary causal inference methods with strict ITT and traditional regression-based mediation analysis. Compared to strict ITT and traditional regression-based mediation analysis, 3 out of 4 of the methods we studied that focus on the controlled direct effect (the QPAC_1.5_, SMM with EE, SMM with g-estimation) resulted in increased magnitude of the treatment effect estimates. In contrast, one method, SMM with GMM, did not increase the size of effect estimates. These findings indicate potential benefits from using these causal inference methods in pain RCTs, but also indicate some instances where they may not perform as expected.

Recent FDA guidance has drawn attention to how intercurrent treatment, such as analgesic use, may affect treatment effect estimates produced by RCTs. [10] “Hypothetical strategies” targeting estimands other than strict ITT have been suggested as alternatives that may better account for intercurrent treatment.[10] However, there have been few studies to date comparing methods for targeting alternative estimands in pain RCTs. [21] In the current study, estimates of the attributable to ESI estimand using the QPAC_1.5_, SMM with EE, and SMM with g-estimation were around 0.1 to 0.2 NRS points larger compared to strict ITT and traditional regression-based mediation analysis estimates. Compared to SMM with EE, SMM with g-estimation and SMM with GMM greatly improved estimation efficiency, as reflected by smaller treatment effect SEs. In general, EE is not recommended since it is less efficient than g-estimation and GMM in that it can yield bigger variance of EE estimator, consistent with findings from the current study. [23] The critical difference between g-estimation and GMM is that g-estimation can only allow for one instrumental variable while GMM allows for multiple instrumental variables. The difficulty of applying g-estimation in practice is to choose an appropriate instrument since difference choices of instruments could lead to different results. In the current study, we examined correlations between potential instruments and analgesic use and then selected the interaction between age and randomization as an instrument since a stronger instrument will typically yield a more stable estimate according to the instrumental variable literature. [12] GMM can incorporate multiple instrumental variables, thereby reducing the arbitrariness in selecting instrumental variables. Although theoretically including more instruments always improves efficiency when the sample size is sufficiently large, GMM might not outperform g-estimation if weak instruments were selected and sample size was not large enough. Indeed, in the current study, compared to the results produced by GMM, results from g-estimation seem more reasonable and consistent with the accepted clinical use of analgesics, which is to reduce pain intensity: g-estimation resulted in non-significant though negative point estimates for *θ*_*M*,0_ suggesting beneficial effects on pain intensity from analgesic use. In contrast, GMM gave positive estimates for *θ*_*M*,0_, suggesting detrimental effects of analgesics on pain intensity. This may also explain why, unlike g-estimation and EE, GMM did not result in larger treatment effects of ESI as compared to strict ITT.

We have reported on the QPAC_1.5_ and its potential ability to mitigate shrinkage in estimated treatment effect sizes due to concurrent analgesic use.[22] The current study differed from our previous report in that, when using the QPAC_1.5,_ this analysis accounted not only for opioid use, but for *all* analgesics (including non-opioid medications). We have also recently reported that the QPAC_1.5_ increased the size of estimated treatment effects in a meta-analysis of interventional pain procedure RCTs. [20] The QPAC_1.5_ is a less flexible method to account for concurrent analgesic use, as compared to SMMs, with potential weaknesses such as assuming the same value for the effect of intercurrent analgesic use on pain intensity, without variability. It was included as a comparator method in the current study not because it is necessarily preferable to other methods to account for intercurrent treatment such as SMMs, but because it may ultimately achieve the same goal as more rigorous causal inference methods. This notion was supported by the current study’s estimates of treatment effects on pain intensity using g-estimation, which were similar to estimates produced using the QPAC_1.5_. Potential advantages which might offset theory-based shortcomings of the QPAC_1.5_ are that it is easy to implement without complex biostatistical expertise; it can be adapted to new pain samples if studies analogous to [22] indicate a different average perceived effect of analgesics in a given target population; and it can be used in the clinical setting for individual patients given that the mathematical correction used in the QPAC_1.5_ is applied at the person level (rather than at the group level).

While theoretical and practical concerns may exist about the methods used to account for concurrent analgesic use in this manuscript, as no single method is currently widely-accepted as best practice, it is generally accepted that not accounting for intercurrent treatment is a potential problem in pragmatic pain RCTs. [9] Design features used in explanatory trials to mitigate concurrent analgesic use, such as strict requirements to use study-provided rescue medication only, may not only limit generalizability but create bias in treatment estimates due to differential participant drop-out. [17] For this reason, lack of current knowledge about which method best accounts for concurrent analgesic use should not prevent the use of such methods in pain RCTs. The current study was the first to systematically compare different contemporary causal inference methods when accounting for intercurrent analgesic use in a pain RCT. It is meant as a demonstration of how these methods generally could be used in any pain RCT. However, it represents only a single step in examining the potential usefulness of such methods to account for intercurrent treatment in pain RCTs. We suggest that analyses using these types of methods be regularly included in publications reporting on pain RCTs so a broad evidence base can be built regarding which methods work best, and how they should be implemented across pain RCTs. Over time, this evidence base will facilitate discussion and eventual agreement about best practices for alternative estimands that can account for intercurrent analgesic use (and perhaps other intercurrent treatments).

Some methodological extensions to the current work can be considered in future studies. First, it is possible to explicitly model effect modification to avoid the no effect-modifier assumption in our SMM approach, however, such models will introduce more parameters to estimate. Second, a rigorous way to construct and select instruments might be useful for further improving the performance of g-estimation and GMM. Third, both g-estimation and GEE may be further improved by using machine learning for the estimation of conditional mean functions. [2]

In conclusion, the current study demonstrates the potential usefulness of contemporary causal inference methods to account for intercurrent analgesic use in in pain RCTs. Future studies are needed to apply similar methods to other pain RCTs to gain expanded experience and inform future analyses.

## Acknowledgements

Research reported in this publication was supported by the University of Washington Clinical Learning, Evidence And Research (CLEAR) Center for Musculoskeletal Research Methodologic and Resource Cores. CLEAR is a Core Center for Clinical Research (CCCR) funded by P30AR072572 from the National Institute of Arthritis and Musculoskeletal and Skin Diseases (NIAMS) of the National Institutes of Health. The content is solely the responsibility of the authors and does not necessarily represent the official views of the National Institutes of Health. Separate from the current work, Dr. Suri is a Staff Physician at the VA Puget Sound Health Care System in Seattle, Washington.

## Data availability

The LESS study data are available for approved internal research purposes under a formal data use agreement. For access inquiries, please contact the CLEAR Center (https://theclearcenter.org/about/resource-core/).

## Supplementary materials to “Estimating controlled direct treatment effects on pain intensity using structural mean models: application to pain randomized controlled trials”

### 1 Notations, causal estimands and assumptions

Throughout this appendix, we will assume our data {*O*_*i*_ = (*X*_*i*_, *R*_*i*_, *M*_*i*_, *Y*_*i*_), *i* = 1, …, *n*} are i.i.d. copies of *O* ∼ *P*_0_. We will drop the subscript *i* when there is no confusion. Here *R* is the randomized treatment assignment (*R* = 1 refers to glucocorticoids plus lidocaine group, *R* = 0 refers to lidocaine alone group). *M* is the binary concurrent analgesics use. *Y* is the outcome measure (LPI or BPI). *X* is a set of baseline covariates. We will adopt the potential outcome framework. Let *Y* (*r, m*) be the potential outcome of *Y* when *R* is fixed at *r* and *M* is fixed at *m, M* (*r*) be the potential outcome of *M* when *R* is fixed at *r*. We will use the notation *Y* (*r*) to denote *Y* (*r, M* (*r*)). Let *U* be unmeasured confounder. We use 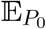 to denote expectation under *P*_0_. We will use empirical process notation *Pf* = ∫ *f* (*o*)*dP* (*o*) for function *f* : *o ↦ f* (*o*). Let *P*_*n*_ be the empirical measure and 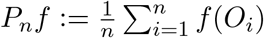 for any function *f*.

We will adopt the following consistency assumption:

#### Assumption 1.

***(Consistency)***. *M* (*r*) = *M, Y* = *Y* (*m, r*) *if R* = *r, M* = *m*.

Since the LESS trial is a randomized clinical trial, throughout this paper we will assume the *R* is independent of any pre-randomization variables and potential outcomes:

#### Assumption 2.

***(Randomization)***. *R ╨* (*X, M* (*r*), *Y* (*m, r*), *U*).

The key estimand in our paper is the controlled indirect effect (CDE) of *R*, that is

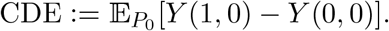

Under the setting of LESS trial, the estimand is also the attributable to ESI estimand. In the supplementary material, we will use the terminology CDE rather than attributable to ESI estimand for broader applications.

### 2 Technical details of intention to treat analysis

The intention to treat analysis is the causal effect of randomized treatment assignment on the outcome, that is:

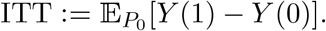

Due to Assumption 2 and Assumption 1, we have

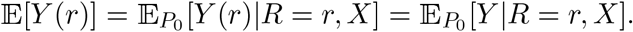

Therefore,

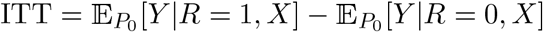

we can fit a regression model of *Y* on *R* and baseline covariates:

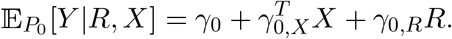

*γ*_0,*R*_ is the ITT. The set of covariates included in the analysis doesn’t affect the unbiasedness of estimated effect but it may affect the efficiency (Imbens and Rubin, 2015).

### 3 Technical details of traditional regression based mediation analysis

The traditional regression-based mediation analysis is based on two nested regression models. The first one is regress *M* on *R* and *X*, the second one is regress *Y* on *M, R* and *X*. Under the assumption that the causal relationship between the mediator and outcome is not confounded by unmeasured covariates and treatment induced covariates, the regression-based mediation analysis yields unbiased estimates for controlled direct and indirect effect Imai et al. (2010). Since here we only care about the direct effect of *R* on *Y*, regressing *Y* on *M, R* and *X* is sufficient for our purpose. To see this, consider the following model

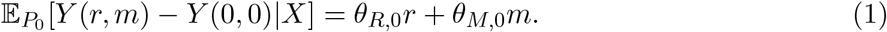

The parameter *θ*_*R*,0_ encodes the direct effect of treatment assignment. *θ*_*M*,0_ encodes the causal effect of analgesic use on the outcome. However, traditional regression methods requires the following two additional assumptions:

#### Assumption 3.

***(Linearity)***. 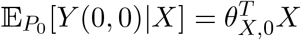.

#### Assumption 4.

***(No unmeasured confounding between*** *M* ***and*** *Y* ). *M* (*r*) ╨ *Y* (*r, m*)|*R* = *r, X*.

Under Assumption 1-4 and model (1), we have the identification of 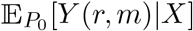:

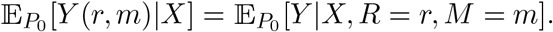

Therefore, regressing *Y* on *X, R* and *M* allows us to estimate the direct effect of *R*.

### 4 Technical details of structural mean models and corresponding estimation strategies

Assumption 3 is a commonly made modelling assumption in practice, however, possible misspecification of *g*_0_(*X*) could induce bias. Assumption 4 is hard to justify in practice. Consider the following structural mean model:

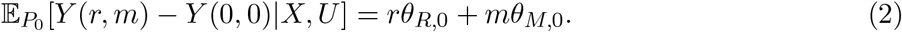

We make the following latent ignorability assumption:

#### Assumption 5.

***(Latent ignorability)***.

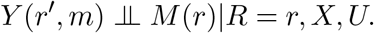

Now we give the formal result on identifiability of *θ*_*R*,0_ and *θ*_*M*,0_.

#### Lemma 1.

*Under Assumptions 1, 2 and 5, assuming model 2 holds*,

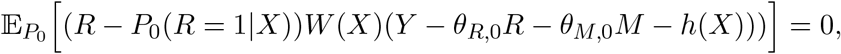

*where W* (*X*) *and h*(*X*) *are any two functions of X*.

*Proof*. Assumptions 1, 2 and 5 together implies

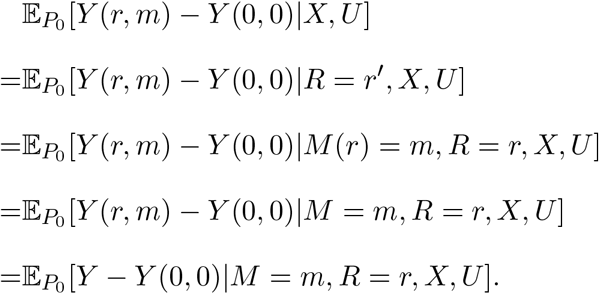

Combing with model 2, we have

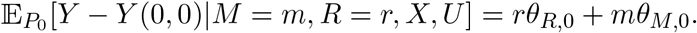

Let 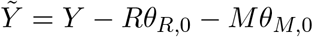, then

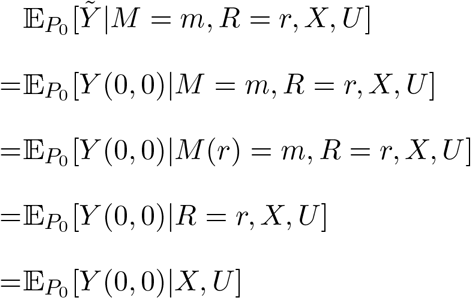

Therefore,

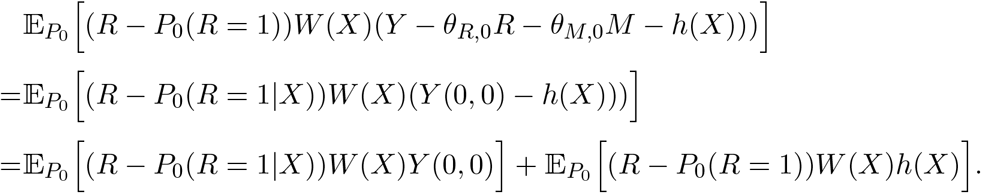

The second term is clearly zero, for the first term,

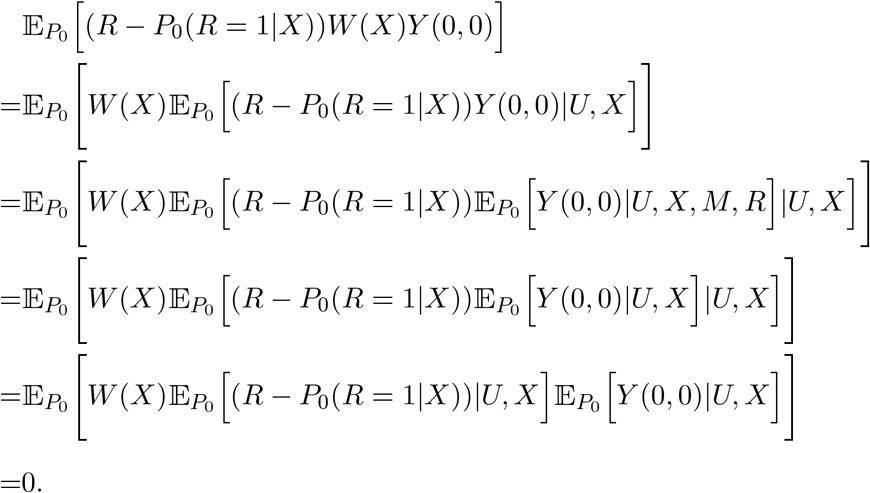

#### 4.1 Estimation equation approach

Since the *h*(*X*) can be any functions of *X*, the first approach is to specify it as zero. Then the estimation equation becomes:

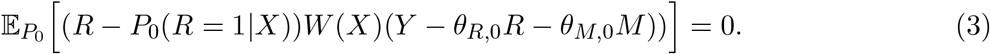

By our study design, we know *P*_0_(*R* = 1|*X*) = *P*_0_(*R* = 1) = 0.5. By specifying *W* (*X*) to be a two dimensional vector, we can solve the empirical version to obtain an estimator 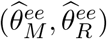 for (*θ*_*R*,0_, *θ*_*M*,0_). That is, 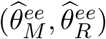 solves the following equation:

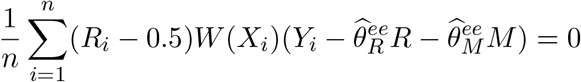

The variance estimator can be obtained using standard estimating equation theory (van der Vaart, 1998). Let

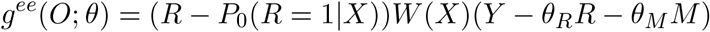

and let

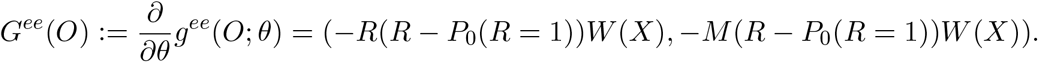

Then the asymptotic variance is

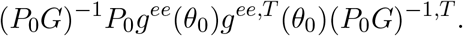

Then the plug-in estimator for asymptotic variance can be constructed as

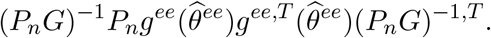

#### 4.2 G-estimation

The estimating equation estimator can be inefficient. Ten Have et al. (2007) described G-estimation procedure for (*θ*_*R*,0_, *θ*_*M*,0_), which involves specifying a linear working model for *h*(*X*).

The g-estimator, denoted as 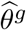, involves iterating the following three steps:

1. Given an estimate 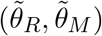, calculate 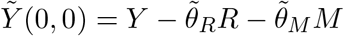.
2. Regress 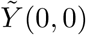 on *X*, obtain an estimate 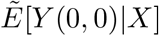.
3. Solve the estimating equation and update the estimate of *θ*_*R*_ and *θ*_*M*_, that is, solving
4. 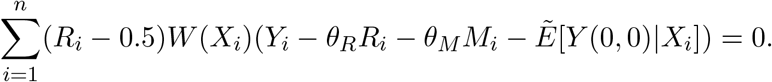

#### 4.3 Generalized method of moments

By orthogonizing the moment condition (3), we can obtain an Neyman orthogonal moment condition (Chernozhukov et al., 2018):

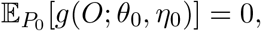

Where

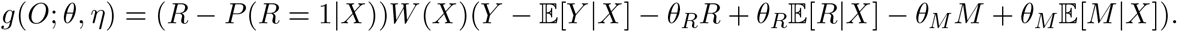

Similar to the estimating equation approach, one can do a two-step estimating equation approach. In the step one, specify models for 𝔼 [*Y*|*X*] and 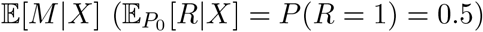.Then plug in the estimated nuisance parameters, denoted as 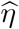, into the estimating equation

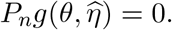

then solve for *θ*. Just like the g-estimation approach, fitting nuisance parameters might improve efficiency.

When *W* (*X*) is more than two dimensional, we can use two-step generalized method of moment (GMM) to estimate *θ*. Including more moment conditions could also improve efficiency. Let *S* be any weighting matrix. The generalized method of moments estimator 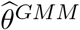 solves the following problem:

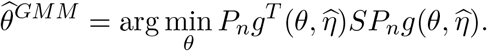

If all the nuisance parameters are belong to correctly specified parametric model, by the Neyman orthogonality property of *g*, the asymptotic variance of 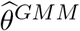 will be the same as the variance of 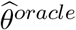, where

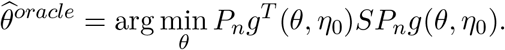

Therefore, the asymptotic variance is

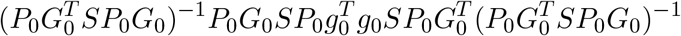

Where

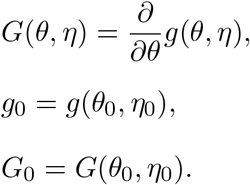

The estimator for asymptotic variance can be constructed as

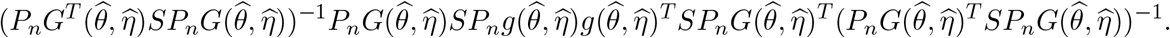

By choosing 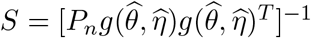, we can obtain the efficient GMM estimator, denoted as 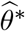.

The over-identification test (J-test) is a test for the null hypothesis: 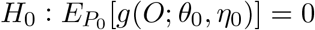. The test statistic can be constructed as

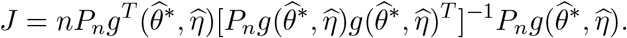

The statistic *J* is asymptotically *χ*^2^(*m* − *p*) distributed under the null hypothesis, where *p* is the dimension of *θ*.

## Reference

[1] Baron RM, Kenny DA. The moderator–mediator variable distinction in social psychological research: Conceptual, strategic, and statistical considerations. Journal of personality and social psychology 1986;51(6):1173.

[2] Chernozhukov V, Chetverikov D, Demirer M, Duflo E, Hansen C, Newey W, Robins J. Double/debiased machine learning for treatment and structural parameters: Oxford University Press Oxford, UK, 2018.

[3] Dworkin RH, Turk DC, Farrar JT, Haythornthwaite JA, Jensen MP, Katz NP, Kerns RD, Stucki G, Allen RR, Bellamy N. Core outcome measures for chronic pain clinical trials: IMMPACT recommendations. pain 2005;113(1):9–19.

[4] FDA. Adjusting for Covariates in Randomized Clinical Trials for Drugs and Biological Products. In: USDoHaH Services, FaD Administration, CfDEaR (CDER), CfBEaR (CBER), OCoE (OCE) editors, 2023.

[5] Friedly JL, Bresnahan BW, Comstock B, Turner JA, Deyo RA, Sullivan SD, Heagerty P, Bauer Z, Nedeljkovic SS, Avins AL. Study Protocol-Lumbar Epidural Steroid Injections for Spinal Stenosis (LESS): a double-blind randomized controlled trial of epidural steroid injections for lumbar spinal stenosis among older adults. BMC musculoskeletal disorders 2012;13:1–9.

[6] Friedly JL, Comstock BA, Turner JA, Heagerty PJ, Deyo RA, Sullivan SD, Bauer Z, Bresnahan BW, Avins AL, Nedeljkovic SS. A randomized trial of epidural glucocorticoid injections for spinal stenosis. New England Journal of Medicine 2014;371(1):11–21.

[7] Hansen LP. Large sample properties of generalized method of moments estimators. Econometrica: Journal of the econometric society 1982:1029–1054.

[8] Hernán M, Robins J. Causal Inference: What If: Boca Raton: Chapman & Hall/CRC, 2020.

[9] Hohenschurz-Schmidt D, Cherkin D, Rice ASC, Dworkin RH, Turk DC, McDermott MP, Bair MJ, DeBar LL, Edwards RR, Evans SR. Methods for pragmatic randomized clinical trials of pain therapies: IMMPACT statement. Pain 2024;165(10):2165–2183.

[10] ICH. Addendum on estimands and sensitivity analysis in clinical trials to the guideline on statistical principles for clinical trials E9(R1), 2021.

[11] Imai K, Keele L, Yamamoto T. Identification, inference and sensitivity analysis for causal mediation effects. 2010.

[12] Imbens G, Rosenbaum P. Robust, accurate confidence intervals with a weak instrument: quarter of birth and education. JOURNAL OF THE ROYAL STATISTICAL SOCIETY SERIES A-STATISTICS IN SOCIETY 2005;168:109–126.

[13] Imbens GW, Rubin DB. Causal inference in statistics, social, and biomedical sciences: Cambridge university press, 2015.

[14] Robins JM. Correcting for non-compliance in randomized trials using structural nested mean models. Communications in Statistics-Theory and methods 1994;23(8):2379–2412.

[15] Robins JM, Greenland S. Identifiability and Exchangeability for Direct and Indirect Effects. Epidemiology 1992;3(2).

[16] Rubin DB. Estimating causal effects of treatments in randomized and nonrandomized studies. Journal of Educational Psychology 1974;66(5):688–701.

[17] Singla NK, Meske DS, Desjardins PJ. Exploring the interplay between rescue drugs, data imputation, and study outcomes: conceptual review and qualitative analysis of an acute pain data set. Pain and Therapy 2017;6:165–175.

[18] Small DS. Mediation analysis without sequential ignorability: using baseline covariates interacted with random assignment as instrumental variables. arXiv preprint arXiv:11091070 2011.

[19] Splawa-Neyman J, Dabrowska DM, Speed TP. On the application of probability theory to agricultural experiments. Essay on principles. Section 9. Statistical Science 1990:465–472.

[20] Sridhar BV, Humbert A, Babitts A, Staab C, Daniels CJ, Dhillon M, Heagerty P, Goldenberg J, Jensen M, Suri P. Methods used to Account for Concurrent Analgesic Use in Randomized Controlled Trials of Interventional Pain Treatments: A Meta-Epidemiologic Study. medRxiv 2024:2024.2011.2002.24316637.

[21] Suri P, Heagerty PJ, Korpak A, Jensen MP, Gold LS, Chan KCG, Timmons A, Friedly J, Jarvik JG, Baraff A. Improving power and accuracy in randomized controlled trials of pain treatments by accounting for concurrent analgesic use. The journal of pain 2023;24(2):332–344.

[22] Suri P, Heagerty PJ, Timmons A, Jensen MP. Description and initial validation of a novel measure of pain intensity: the Numeric Rating Scale of Underlying Pain without concurrent Analgesic use. Pain 2024;165(7):1482–1492.

[23] Ten Have TR, Joffe MM, Lynch KG, Brown GK, Maisto SA, Beck AT. Causal mediation analyses with rank preserving models. Biometrics 2007;63(3):926–934.

[24] Van der Vaart AW. Asymptotic statistics, Vol. 3: Cambridge university press, 2000.

[25] Vansteelandt S, Joffe M. Structural nested models and G-estimation: the partially realized promise. 2014.

[26] White H. A heteroskedasticity-consistent covariance matrix estimator and a direct test for heteroskedasticity. Econometrica: journal of the Econometric Society 1980:817–838.

## References

Chernozhukov, V., Chetverikov, D., Demirer, M., Duflo, E., Hansen, C., Newey, W., and Robins, J. (2018). Double/debiased machine learning for treatment and structural parameters. The Econometrics Journal, 21(1):C1–C68.

Imai, K., Keele, L., and Yamamoto, T. (2010). Identification, Inference and Sensitivity Analysis for Causal Mediation Effects. Statistical Science, 25(1):51 – 71.

Imbens, G. W. and Rubin, D. B. (2015). Causal inference in statistics, social, and biomedical sciences. Cambridge university press.

Ten Have, T. R., Joffe, M. M., Lynch, K. G., Brown, G. K., Maisto, S. A., and Beck, A. T. (2007). Causal mediation analyses with rank preserving models. Biometrics, 63(3):926–934.

van der Vaart, A. W. (1998). Asymptotic Statistics. Cambridge Series in Statistical and Probabilistic Mathematics. Cambridge University Press.

